# Integrating AI-powered automated neurovascular bundle segmentation and radiomics for prostate cancer staging

**DOI:** 10.64898/2026.03.10.26347880

**Authors:** Gemma Urbanos, Anna Nogué-Infante, Gloria Ribas, Francisco Higa, Marialaura Mena-Clavelis, Claudia Fontenla-Martínez, Polina Rudenko, Eduardo Baettig, Vicente Belloch-Ripollés, Almudena Fuster-Matanzo, Luis Martí-Bonmatí, Angel Alberich-Bayarri, Ana Jimenez-Pastor

## Abstract

**Background:** Assessment of the prostatic neurovascular bundles on MRI is clinically relevant for staging and treatment planning but remains technically challenging and underexplored in automated imaging pipelines.

**Purpose:** To develop and evaluate an automated framework for neurovascular bundle segmentation, proximity-based invasion risk stratification, and radiomics-based prediction of biochemical recurrence, perineural invasion, and extraprostatic extension.

**Methods:** This retrospective study included 808 prostate MRI examinations from three datasets acquired between 2015 and 2020. Among them, 470 T2-weighted image sequences were manually annotated to train a 3D full-resolution nnU-Net segmentation model. Tumor-to-neurovascular bundle distance was used to define invasion risk categories (low, intermediate, high). Machine learning models were developed using radiomics features extracted from neurovascular bundles, lesions, and combined regions, with optional inclusion of prostate-specific antigen and age at MRI. Model performance was evaluated using area under the receiver operating characteristic curve and accuracy. Model interpretability was assessed using Shapley additive explanations.

**Results:** The median patient age was 69 years (interquartile range, 63–73). Automatic neurovascular bundles segmentation achieved anatomically plausible contours, with average surface distance below 1 mm and volume difference under 0.4 cc. The resulting tumor-to-neurovascular bundle invasion risk classification reached 90% accuracy, supporting usability. Radiomics models showed predictive value across endpoints, with moderate testing performance for biochemical recurrence (AUC = 0.73), and higher discrimination for perineural invasion (AUC = 0.80) and extraprostatic extension (AUC = 0.80). Interpretability analysis revealed that tumor-to-neurovascular bundle proximity and neurovascular bundles imaging features among the most relevant contributors to outcome prediction.

**Conclusions:** Automated neurovascular bundle segmentation enabled quantitative tumor proximity assessment and radiomics-based prediction of biochemical recurrence, perineural invasion, and extraprostatic extension.

## Introduction

Prostate cancer (PCa) is one of the most prevalent malignancies worldwide and the most frequently diagnosed solid tumor in men^1^. Although multiparametric MRI (mpMRI) is the reference imaging standard, biparametric MRI (bpMRI) has gained acceptance due to reduced acquisition time and cost while maintaining non-inferior diagnostic accuracy for clinically significant disease^2^. Despite often being diagnosed at a localized stage, PCa shows marked biological heterogeneity, making accurate assessment of local tumor extent critical for personalized treatment planning.

Neurovascular bundle (NVB) involvement is a key factor for nerve-sparing surgery, radiotherapy, and conservative management^3^. Located posterolaterally to the prostate, NVBs contain neural and vascular structures essential for erectile and sphincter function^4^, and their invasion is associated with adverse pathological features and higher surgical margin risk. However, reliable MRI assessment remains challenging due to their small size, anatomical variability, and limited contrast^5,6^. Recent deep learning–based automatic segmentation approaches offer a promising solution to improve consistency and enable systematic extraction of radiomics features from tumors and peri-NVB regions^7^. Radiomics has shown value for predicting tumor aggressiveness and clinical outcomes in PCa^8,9^, while peri-NVB descriptors may capture imaging correlates of perineural invasion and extraprostatic extension^10,11^.

We hypothesized that automatic segmentation of the neurovascular bundles enables objective and reproducible quantification of tumor–NVB spatial relationships on MRI, providing a framework for the extraction of imaging biomarkers with potential diagnostic and prognostic relevance.

Accordingly, this study aimed to (i) develop and validate an automatic NVB segmentation model on T2w MRI, including a proximity-based tumor–NVB risk score, and (ii) assess whether spatial tumor–NVB relationships combined with radiomics and clinical variables can predict diagnostic (PNI, EPE) and prognostic (BCR) outcomes.

## Materials and methods

### Patient cohort and imaging data

#### Study population

This retrospective study cohort comprised 808 prostate MRI examinations from three sources. A total of 537 scans from 366 patients, including diagnostic and follow-up scans acquired between 2015 and 2020, were obtained from an institutional research project. Additionally, 74 examinations were retrieved from multicenter European research initiative^12^, 83 scans were collected from an academic medical center in the United States, and 99 were acquired from a commercial medical imaging repository. Finally, the dataset was complemented with 15 publicly available scans with neurovascular bundle annotations from The Cancer Imaging Archive (TCIA)^13^. The institutional cohort comprised consecutive patients meeting the inclusion criteria during the study period. All data were collected under observational research protocols approved by the corresponding local Ethics Committees, with a waiver of informed consent. Figure 1 illustrates the number of examinations included at each stage of the study workflow.

**Figure 1.**
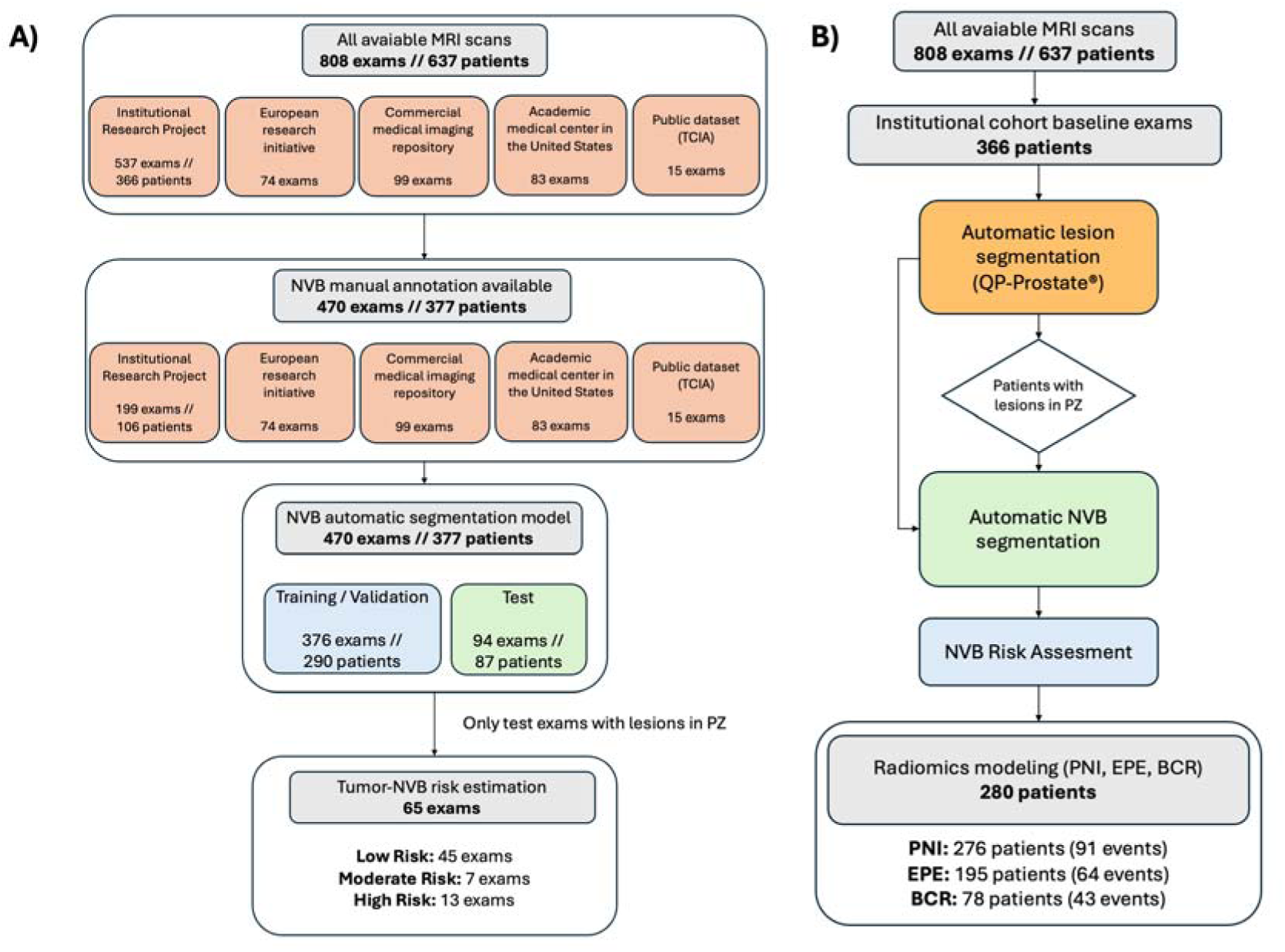
Patient flow across the different stages of the study workflow: (A) the NVB segmentation and invasion risk assessment pipeline and (B) the radiomics analysis workflow. NVB, neurovascular bundle.

Eligible patients had biopsy-proven PCa with Gleason score ≥ 6 and available bpMRI (T2w and DWI series). Axial T2w images were selected as the analyzed sequence due to their central role in PCa staging and superior depiction of the prostate and periprostatic NVBs^14^. Exclusion criteria included prior prostatectomy, severe imaging artifacts, and fat-suppressed T2w acquisitions, which were applied during dataset curation to ensure imaging homogeneity. All included examinations had complete imaging, annotation, and clinical data required for the analyses, and no missing data handling was necessary.

### Imaging data and annotations

All 808 MRI studies were processed and annotated using QP-Insights® (Quibim S.L., Spain). DICOM headers were pseudonymized prior to analysis using project-specific identifiers to enable linkage with clinical variables. The bpMRI series were used for prostate gland and intraprostatic automatic lesion segmentation with QP-Prostate® (Quibim S.L., Spain). Manual slice-by-slice delineation of the NVBs was performed on the axial T2w images by expert readers (four radiologists and three Quibim trained imaging technicians), focusing on NVB segments at the prostatic base where these structures are most reliably visualized. Annotators were blinded to model predictions and clinical outcomes during manual segmentation.

The final annotated dataset comprised 470 exams from 377 patients. Representative examples illustrating NVB anatomical variability and indistinct borders are shown in Figure 2. Manual expert delineation was used as the reference standard, as expert segmentation is the accepted ground truth for anatomical structure definition in medical image segmentation, particularly in the absence of spatially matched histopathology.

**Figure 2.**
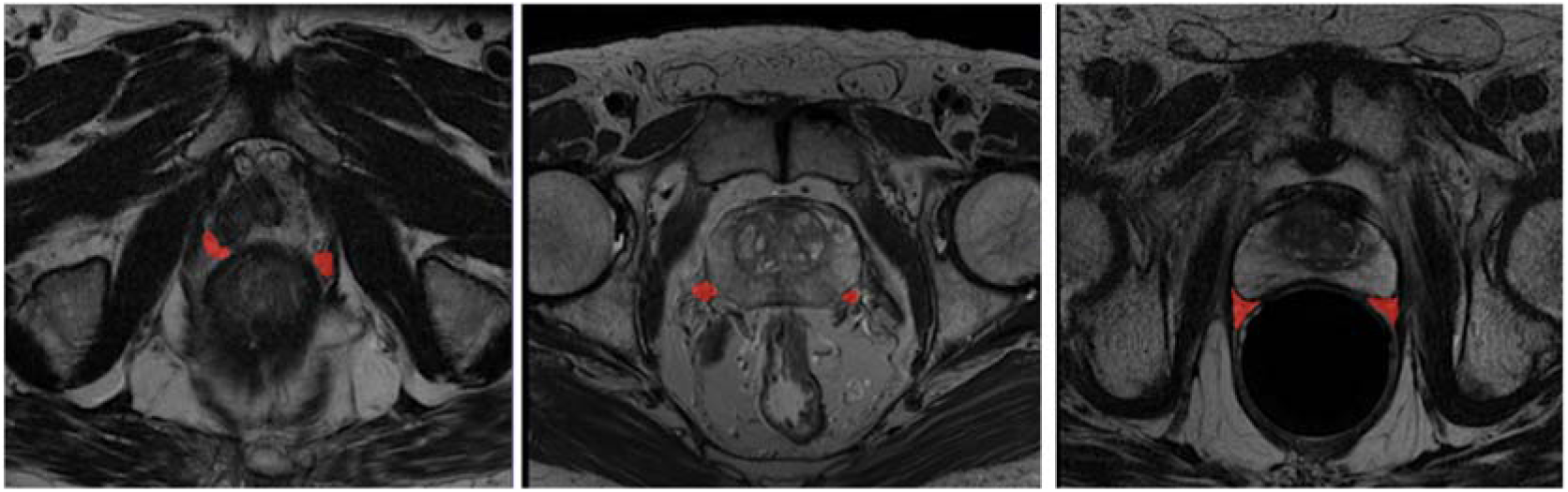
Examples of NVB manual segmentations. NVB, neurovascular bundle.

### Neurovascular bundles automatic segmentation

A total of 470 annotated exams were included. Within each dataset, cases were randomly divided into 80% for model development and 20% for testing, resulting in 376 and 94 exams, respectively. Institutional stratification and patient-level grouping of longitudinal studies were applied to prevent information leakage. Preprocessing included isotropic resampling (1mm^3^), removal of voxels segmented as prostate gland from NVB masks to avoid label overlap, z-cropping to prostate-containing slices, and z-score intensity normalization of T2w images. A nnU-Net 3D full-resolution configuration^15^ was trained for NVB segmentation using T2w images and prostate masks as inputs. Models were trained with five-fold cross-validation and ensembled by averaging voxel-wise probability maps (Supplementary Figure S1).

Performance was evaluated on the test set using Dice similarity coefficient (DSC), sensitivity, average surface distance (ASD), and volume difference^16^, complemented by Bland–Altman analysis^17^ for volumes comparison. DSC was computed using both hard predictions, applying the standard probability threshold of 0.5, and soft formulations derived from continuous probability maps. Test-time augmentation (10 perturbations per exam) was applied and averaged with the original prediction. For qualitative assessment, 50 randomly selected cases were independently reviewed by experienced genitourinary radiologists blinded to the reference standard annotations.

Robustness analyses stratified performance by clinically relevant factors, including benign prostatic hyperplasia (BPH)^18^, high-risk NVB invasion (tumor-to-NVB distance < 2 mm^19^), and the presence of peripheral-zone (PZ) lesions. Statistical testing was adapted to data distribution, assessed using Shapiro–Wilk and Levene’s tests, with parametric (Student’s or Welch’s t-test) or non-parametric (Mann–Whitney U) comparisons applied as appropriate; significance was set at p ≤ 0.05.

### Tumor-to-NVB risk estimation

To assess the clinical utility of the segmentation model, the spatial relationship between prostate tumors and the NVBs was evaluated in the test set (94 exams). Patients without visible tumors and lesions confined to the transition and central zones were excluded from NVB-related analyses (65 exams). For the remaining exams, the 3D minimum Euclidean distance between each lesion and both NVBs was computed using manual ground-truth and model-predicted masks. For patients with multiple lesions, the overall risk category corresponded to the lesion closest to a NVB.

Tumor-to-NVB proximity was subsequently stratified into three predefined clinical risk categories: high (<2 mm), intermediate (2–5 mm), and low (>5 mm). Threshold selection was guided by prior evidence: anatomical studies place the NVBs within roughly 5 mm of the prostatic capsule^6^, and distances greater than 2 mm are generally considered safe for nerve preservation during surgery^19^.

### Clinical relevance of segmentation-derived features

Clinically relevant outcomes (EPE, PNI, and BCR) were used as prediction targets. MRI served as the index test, while clinical outcomes obtained from histopathology and follow-up records served as the reference standard. EPE reflects extracapsular tumor extension and its relationship with NVB proximity, PNI captures tumor spread along neural structures, and BCR represents downstream tumor aggressiveness, for which lesion and NVB radiomics may provide complementary prognostic information.

Models were developed using MRI radiomics and clinical variables available at diagnosis (PSA and age), enabling outcome prediction from the diagnostic time point. The complete analytical pipeline is summarized in Figure 3.

**Figure 3.**
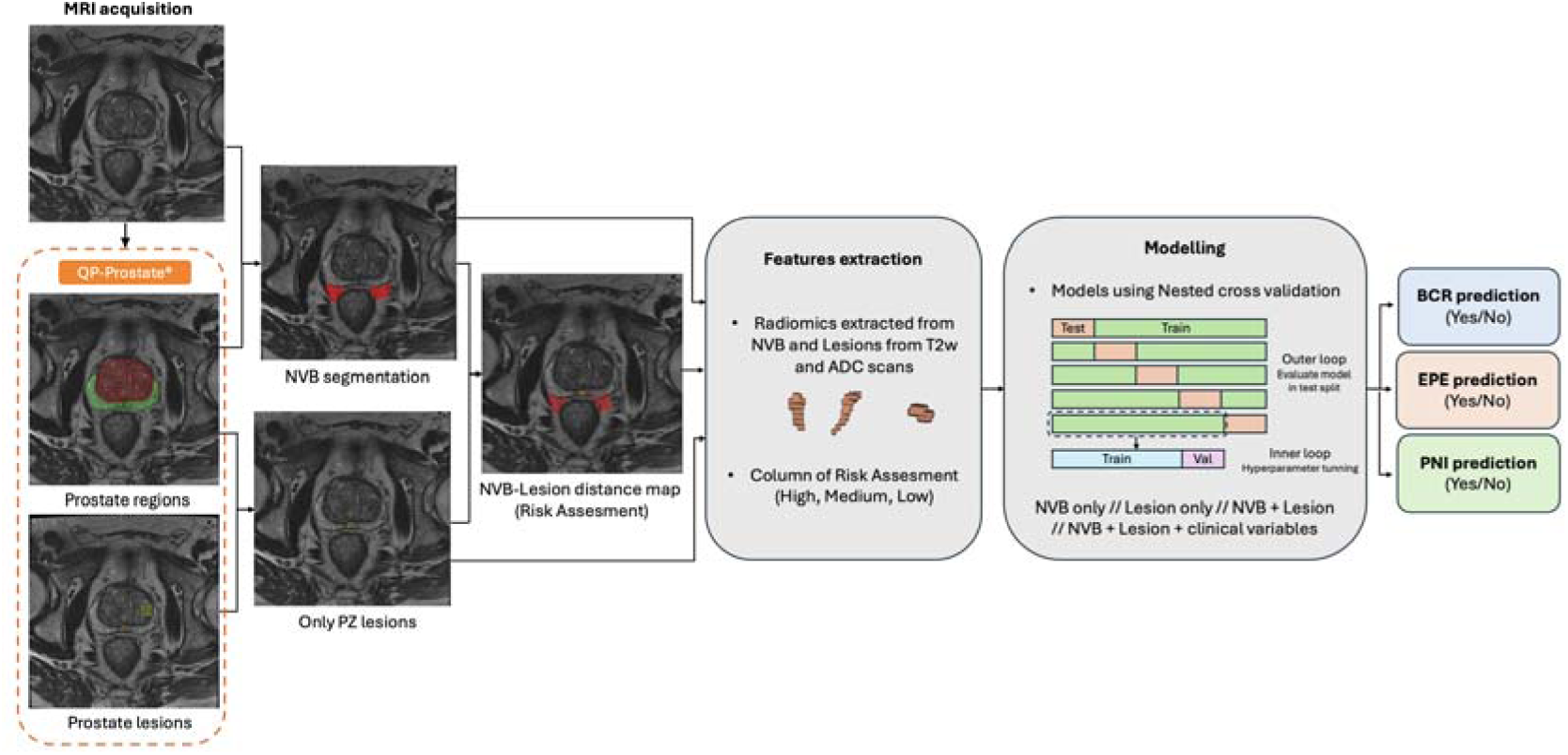
Pipeline for outcome prediction using radiomics features from lesions, NVBs segmentations and clinical data. MRI, magnetic resonance imaging; NVB, neurovascular bundles; PZ, peripheral zone; T2w, T2-weighted; ADC, apparent diffusion coefficient; BCR, biochemical recurrence; EPE, extraprostatic extension; PNI, perineural invasion.

### Radiomics feature extraction and evaluated feature set design

Radiomic analysis quantified imaging-derived biomarkers from prostate lesions and NVBs. Features were extracted from T2w images and apparent diffusion coefficient (ADC) maps computed from DWI using QP-Prostate^®^ module within QP-Insights^®^ platform yielding 1,379 features per region of interest (ROI), including shape, first-order intensity, texture, and wavelet-and Laplacian of Gaussian–based features.

To assess the incremental value of anatomical regions and data modalities, four model configurations were evaluated: (i) NVB radiomics combined with NVB invasion risk, (ii) lesion radiomics combined with NVB invasion risk, (iii) combined lesion and NVB radiomics, and (iv) multimodal models integrating lesion and NVB radiomics with NVB invasion risk and clinical variables (PSA and age).

### Radiomics-based machine learning model

Categorical variables were encoded for model integration, with NVB invasion risk treated as an ordinal variable (high, intermediate, low), and all continuous features standardized using z-score normalization. The machine learning pipeline followed a nested 5×5 cross-validation scheme, with performance reported as the mean across outer test folds. Random forest, extra trees, and XGBoost models were evaluated using AUC as the primary metric. Performance metrics were calculated across cross-validation folds, and mean values with corresponding 95% confidence intervals were reported.

Feature redundancy was controlled using Spearman correlation, removing variables with |ρ| > 0.90 by discarding one feature per correlated pair. Z-score normalization was applied within each fold. Models were trained with and without outlier removal using Isolation Forest^20^, and feature selection was performed using maximum relevance minimum redundancy (MRMR) with 10, 20, or 30 features^21^. Class imbalance was addressed through combined over- and under-sampling strategies^22^. Hyperparameters were optimized via grid search (Supplementary Table S1).

### Clinical outcome definitions

The clinical outcomes considered for prediction were EPE, PNI, and BCR:

- EPE: tumor extension beyond the prostatic capsule on histopathology, encoded as a binary variable (0 = absent, 1 = present)^23^.
- PNI: histological evidence of tumor cells infiltrating or tracking along periprostatic nerves, also coded as binary (0 = absent, 1 = present)^24^.
- BCR: defined as PSA ≥ 0.2 ng/mL after prostatectomy (confirmed by a second test), or nadir PSA + 2 ng/mL after radiotherapy according to the Phoenix criteria; modeled as a binary outcome (0 = no recurrence, 1 = recurrence)^25^.

### Model interpretability

Model interpretability was assessed using Shapley additive explanations (SHAP)^26^. SHAP values from the best cross-validated model on the test set quantified feature contributions, with global analyses identifying influential radiomics and clinical variables and local explanations illustrating case-level effects.

## Results

### Cohort selection and outcome-driven database characterization

The study integrated three complementary datasets with heterogeneous scanner manufacturers, field strengths, and acquisition protocols. The institutional cohort constituted the largest dataset and was mainly acquired on GE 3T systems, whereas the multicenter European initiative, the academic U.S. center, and the commercial repository included mixed Siemens, Philips, and GE scanners, predominantly at 3T. The public dataset was the most homogeneous cohort, comprising exclusively Philips 1.5T acquisitions. Despite this variability, pixel spacing and slice thickness were largely comparable across datasets (Supplementary Table S2).

In the independent test set (n = 94), 65 exams presented PZ lesions, stratified as low (n = 45), intermediate (n = 7), and high NVB invasion risk (n = 13). Radiomics analyses were restricted to institutional cohort patients with PZ tumors and complete outcome data (n = 280). Outcome-specific filtering yielded varying sample sizes and event rates: PNI (n = 276; 91 events), EPE (n = 195; 64 events), and BCR (n = 78; 43 events). The participant flow diagram, detailing the allocation of cases to the training and test sets throughout the study workflow, is shown in Figure 1. Baseline demographic characteristics of the study population are summarized in Table 1. Median age was 69 years (IQR 63–73).

### NVB segmentation performance

On the test set (n = 94), NVB segmentation achieved a median (IQR) hard DSC of 0.61 (0.52–0.65), which increased to 0.66 (0.58–0.70) when using soft DSC from continuous probability maps (Table 2). Spatial errors were limited (ASD=1.02 mm; Hausdorff=16.34 mm), with a sensitivity of 0.55. Bland–Altman analysis of NVB volumes (Supplementary Figure S2) showed minimal bias, with most observations within the 95% limits of agreement and no evidence of proportional bias, although variability increased for larger volumes.

**Table 2.** Performance metrics of the automatic NVB segmentation model in test set (N = 94).

| Metric | Both NVBs | Left NVB | Right NVB |
| --- | --- | --- | --- |
| Total volume (cc) | 2.94 (2.42–3.61) | 1.50 (1.14–1.96) | 1.37 (0.98–1.75) |
| Volume difference (cc) | 0.41 (0.23–0.76) | 0.48 (0.23–0.74) | 0.34 (0.19–0.70) |
| Distance between inferior and superior bounds (mm) | 16.47 (14.72–18.77) | 14.78 (11.91–16.17) | 14.63 (12.03–16.27) |
| Dice score (threshold 0.5) | 0.61 (0.52–0.65) | 0.59 (0.49–0.67) | 0.61 (0.51–0.68) |
| Soft Dice score (probability maps) | 0.66 (0.58–0.70) | 0.57 (0.48–0.67) | 0.56 (0.49–0.68) |
| Average surface distance (mm) | 1.02 (0.58–1.93) | 1.09 (0.60–2.26) | 0.81 (0.46–1.62) |
| Hausdorff distance (mm) | 16.34 (12.20–20.81) | 13.81 (9.90–18.95) | 13.47 (9.49–17.91) |
| Recall | 0.55 (0.47–0.71) | 0.60 (0.47–0.73) | 0.58 (0.43–0.72) |

Visual assessment of representative cases (Figure 4) and slice-by-slice review of 50 randomly selected test cases by experienced genitourinary radiologists confirmed anatomically plausible and consistent contours, accurately following the expected posterolateral NVB course without implausible false positives or missed bundles.

**Figure 4.**
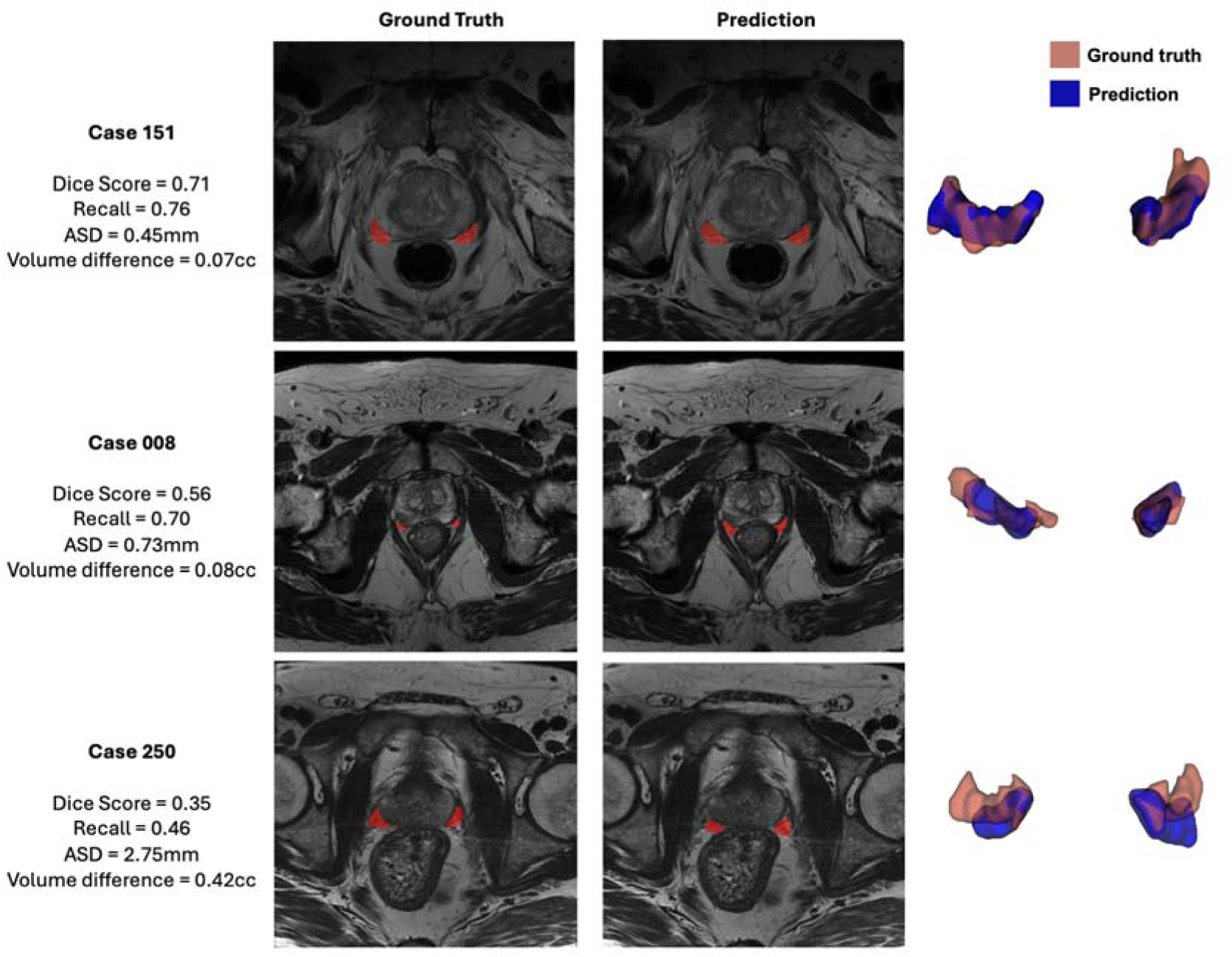
Examples of automatic NVB segmentation (right) compared with the corresponding manual ground truth (left) in the test set. The last column illustrates 3D surface renderings with overlaid ground truth and predicted NVB segmentations. NVB, neurovascular bundle; ASD, average surface distance.

Subgroup analyses across clinically relevant scenarios, including BPH, high NVB invasion risk, and PZ lesion presence, revealed no significant differences in DSC or ASD (Supplementary Table S3), supporting the robustness of the segmentation model.

### Risk score prediction based on tumor distance to NVBs

Table 3 summarizes the risk classification metrics for the left and right NVBs (N=65). The model achieved high overall performance, with a mean precision of 89.4%, recall of 85.8%, and F1-score of 89.0%. Results were comparable for the two sides, although classification of the left NVB was slightly superior. Performance was higher for low- and high-risk categories (F1-scores close to or above 90%), whereas the intermediate-risk category showed reduced performance (close to 70% F1-score), primarily due to the inherent difficulty in accurately distinguishing distances within the narrow 2–5 mm range.

**Table 3.** Invasion risk classification metrics for the left and right NVBs (N=65)

| NVB Laterality | Risk Category | Precision | Recall | F1-Score |
| --- | --- | --- | --- | --- |
| Left | Low risk | 96.89% | 100.00% | 98.00% |
|  | Moderate risk | 100.00% | 60.00% | 75.00% |
|  | High risk | 90.00% | 100.00% | 94.74% |
|  | **Average** | 95.63% | 86.66% | 89.25% |
| Right | Low risk | 94.44% | 94.44% | 94.44% |
|  | Moderate risk | 75.00% | 60.00% | 66.67% |
|  | High risk | 80.00% | 100.00% | 88.89% |
|  | **Average** | 83.15% | 84.81% | 83.34% |

### Prediction of clinically relevant outcomes

Figure 5 summarizes the predictive performance of four radiomics models (Lesion, NVB, Lesion + NVB, and Lesion + NVB + Clinical) across BCR, EPE, and PNI. Overall, the combined Lesion + NVB model achieved the best performance, reaching mean AUCs of 0.73 for BCR, 0.80, for EPE, and 0.80 for PNI, with balanced accuracies of 0.63, 0.72, and 0.71, respectively. The Lesion-only model outperformed NVB-only but remained below those combined approaches. Incorporating clinical variables did not yield clear improvements compared with imaging-based models alone. By outcome, PNI showed the highest discrimination and recall (0.79–0.82), EPE achieved intermediate AUCs with the highest specificity (up to 0.75), while BCR was the most challenging, with lower AUCs, reduced specificity, and wider intervals. Across all outcomes, recall consistently exceeded specificity.

**Figure 5.**
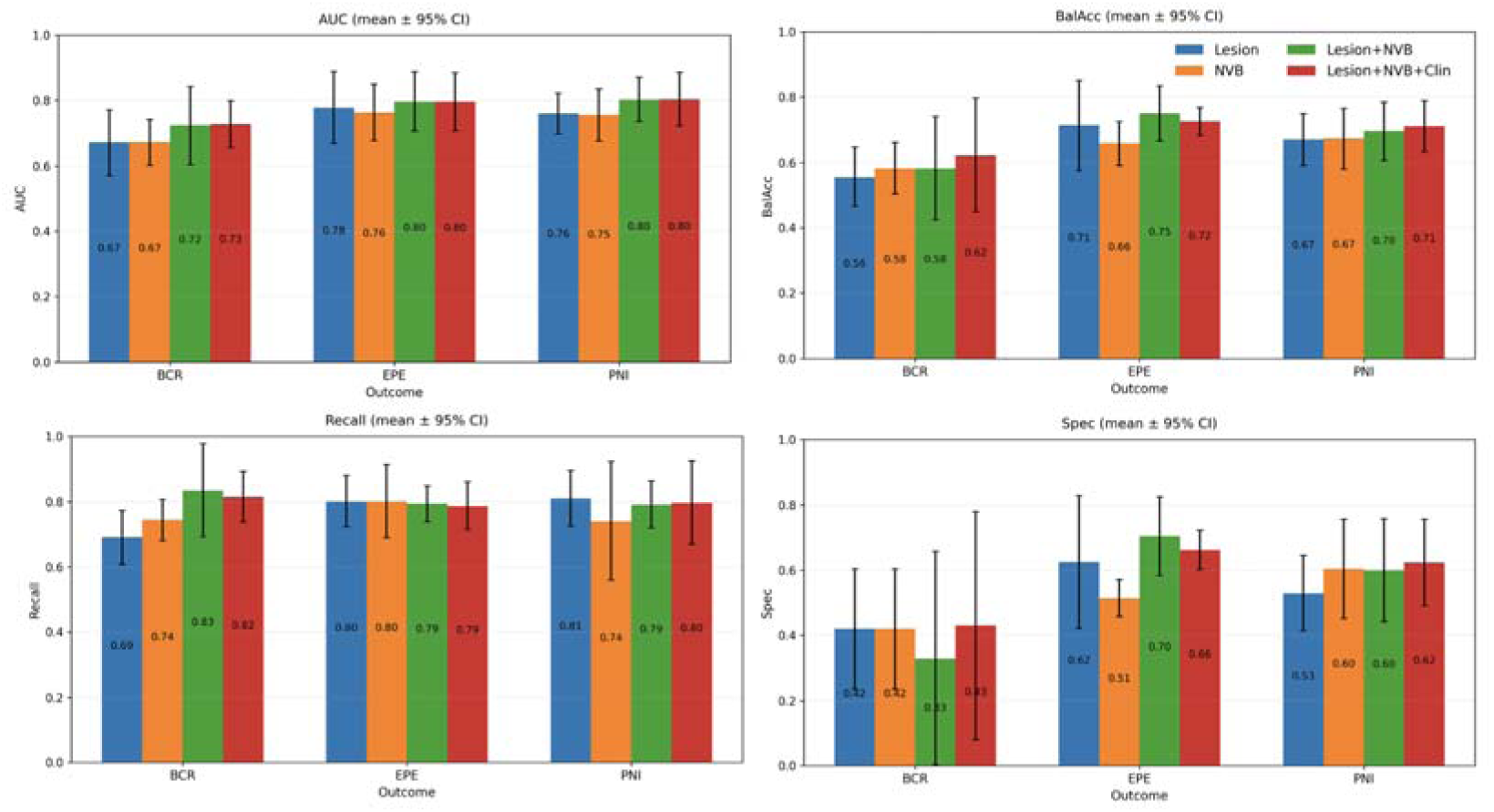
Performance of radiomics models by feature set and outcome. Bars show mean (± 95% CI) for AUC (top left), balanced accuracy (top right), recall (bottom left), and specificity (bottom right). AUC, area under the curve; BalAcc, balanced accuracy; BCR, biochemical recurrence; Clin, clinical variables; EPE, extraprostatic extension; NVB, neurovascular bundles; PNI, perineural invasion; Spec, specificity.

### Interpretability analysis

Task-specific features contributions across PNI, EPE, and BCR. PNI were mainly driven by NVB invasion risk, supported by NVB radiomics and lesion features (Figure 6A). EPE prediction relied primarily on lesion and NVB morphology, with additional contributions from NVB heterogeneity features and PSA (Figure 6B). In contrast, BCR was dominated by NVB texture descriptors, with lesion shape and PSA playing secondary roles (Figure 6C).

**Figure 6.**
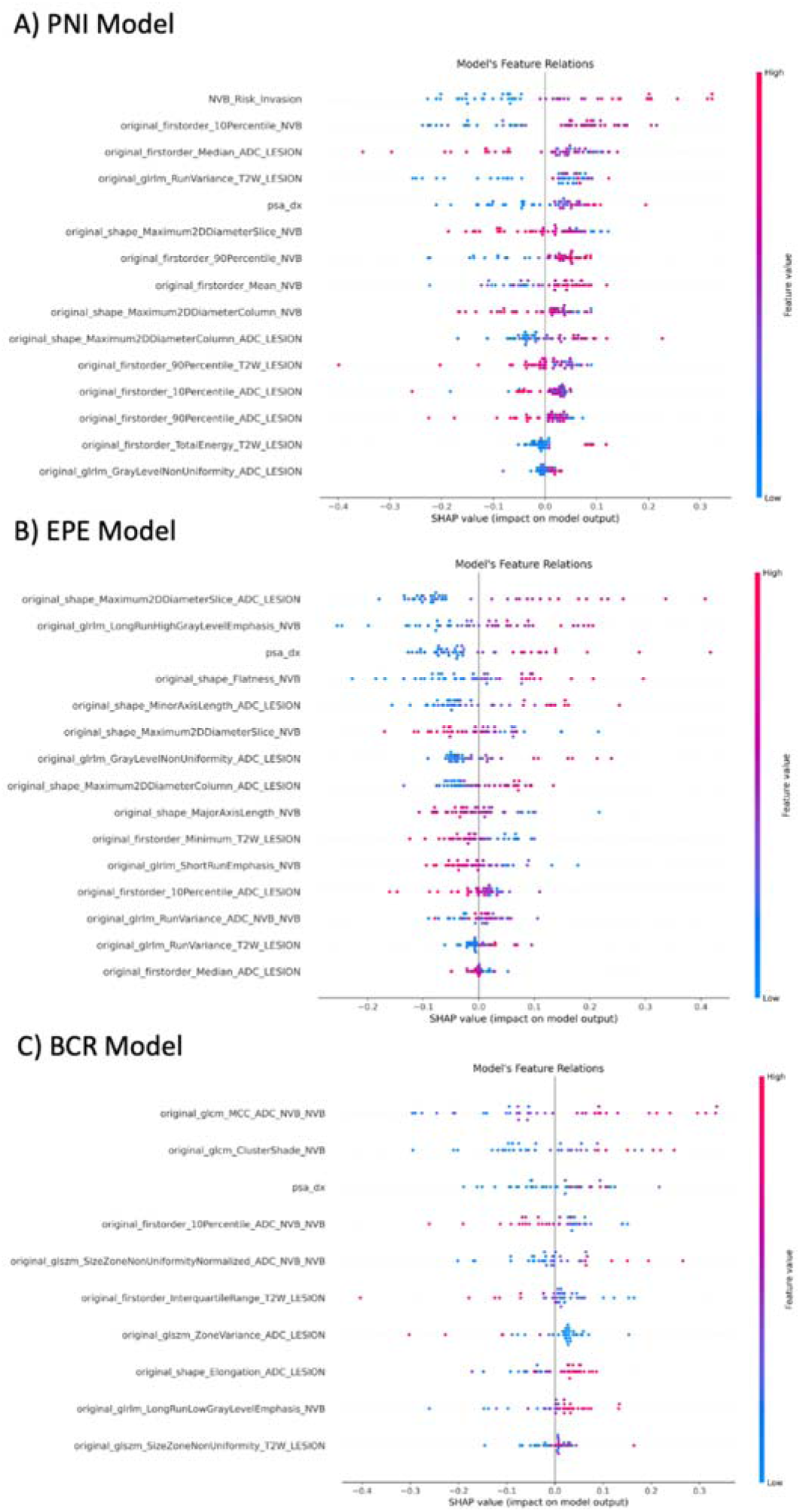
SHAP summary plots showing the relative contribution of clinical and radiomics features to model predictions for PNI, EPE, and BCR. BCR, biochemical recurrence; EPE, extraprostatic extension; PNI, perineural invasion.

## Discussion

Accurate assessment of the prostatic neurovascular bundles (NVBs) is clinically relevant for nerve-sparing treatment planning and outcome prediction but remains challenging due to their small size and high interobserver variability. In this study, we developed a fully automated pipeline for NVB segmentation, tumor–nerve proximity–based invasion risk estimation, and outcome-specific radiomics modeling, addressing a small yet clinically critical anatomical structure. This framework provides a reproducible alternative to expert visual assessment and may support more consistent nerve-sparing surgical and radiotherapy planning. In the test set, NVB segmentation achieved anatomically consistent results (median DSC 0.61; soft DSC 0.66), low spatial errors (ASD 1.02 mm; Hausdorff 16.34 mm), and minimal Bland–Altman bias, enabling reliable quantitative proximity assessment.

Segmentation performance was consistent with prior work. Teunissen et al. reported interobserver variability in manual NVB delineation among four radiation oncologists^27^, and our results fall within these ranges despite relying on annotations from seven raters (Supplementary Table S4). While Teunissen et al. segmented the full neurovascular network, we focused on inferior NVB segments most relevant for nerve-sparing surgery, leading to smaller volumes and increased sensitivity of overlap metrics. Comparable DSC and low surface distances under these stricter conditions support the robustness of the approach.

Left and right NVBs showed highly consistent performance, with minimal differences likely reflecting individual variability rather than model bias. Qualitative review confirmed anatomically plausible contours without unrealistic false positives or missed bundles. Performance remained stable across BPH, NVB invasion status, and PZ lesion presence, indicating reliable behavior under heterogeneous clinical conditions.

Integrating lesion and NVB based radiomics yielded AUCs of 0.73 for BCR, 0.80 for EPE, and 0.80 for PNI. Previous MRI radiomics studies, largely relying on lesion-centric or whole-gland features, have reported AUCs of approximately 0.80 for EPE^28^, ≥0.72 for BCR^29^, and up to 0.88 for PNI^30^. Despite limited cross-study comparability, our results support the added value of NVB radiomics and, to our knowledge, constitute the first integration of automated NVB segmentation into radiomics-based risk stratification using routine pre-treatment bpMRI.

This study has limitations. First, segmentation performance remained moderate, reflecting the anatomical complexity and small size of NVBs, and segmentation uncertainty may propagate into proximity-based features. Second, the relatively limited biochemical recurrence sample size resulted in wider confidence intervals and may affect statistical precision. Third, the single-center design may limit generalizability, and external multicenter validation is required to assess robustness across scanners, acquisition protocols, and patient populations.

In conclusion, this study demonstrates the feasibility and clinical relevance of an end-to-end framework for automated NVB segmentation and the extraction of spatially grounded radiomics biomarkers in PCa. To our knowledge, this is among the first studies to systematically quantify NVB features and to evaluate their contribution to major clinical outcomes. The proposed pipeline, from segmentation to proximity-based risk assessment, provides a robust and reproducible approach that reduces interobserver variability while extending the clinical utility of imaging biomarkers. It generates anatomically consistent NVB contours, minimizes operator-dependent bias, and enables reliable and clinically interpretable invasion-risk stratification that directly supports nerve-sparing surgical and radiotherapy planning. The results achieved have strong potential to enhance diagnostic precision and to enable more individualized treatment strategies.

## Data Availability

All data produced in the present study are available upon reasonable request to the authors

