## Supplementary Material for "Integrating AI-powered automated neurovascular bundle segmentation and radiomics for prostate cancer staging"

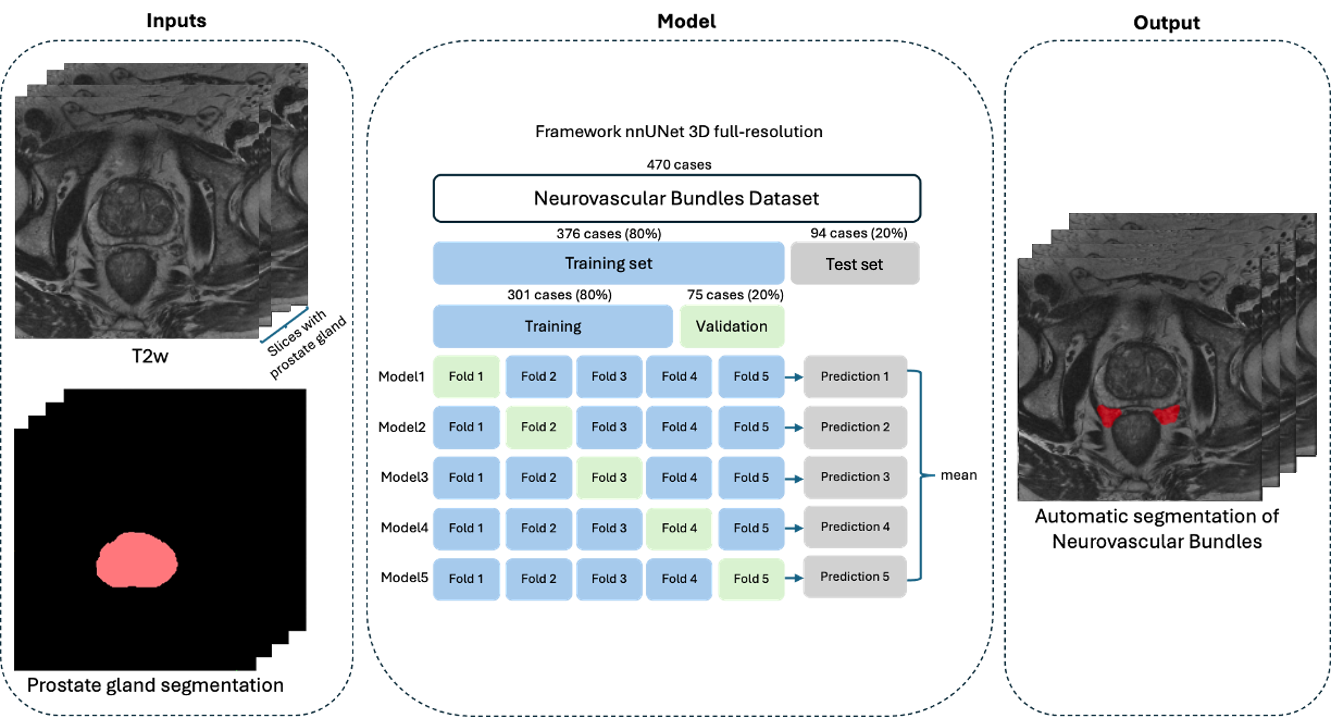

**Figure S1.** Overview of the methodological pipeline. Inputs included T2-weighted and prostate gland segmentation; model development followed a five-fold cross-validation strategy, and the resulting models were ensembled. Final performance was evaluated exclusively on the independent test set.

**Table S1**. Hyperparameter search space for the evaluated classifiers.

| **Classifier** | **Hyperparameters explored** | **Search space** |
| --- | --- | --- |
| RF | Number of trees | {100, 200, 300} |
|  | Maximum depth | {2, 3} |
|  | Min. samples split | {3, 4, 5} |
|  | Min. samples leaf | {2, 3, 4} |
|  | Max. features | {0.15, 0.3, 0.5} |
|  | Split criterion | {gini, entropy} |
| ExtraRF | Same as RF | Same as RF |
| XGBoost | Learning rate | 10 values from 0.001 to 0.15 |
|  | Gamma | 5 values from 10 to 20 |

**Table S2.** Summary of cases and MRI acquisition parameters across cohorts (*N* = 808). NVB, neurovascular bundle.

| **Parameter** | **Category** | **Institutional cohort (n = 537)** | **Externals cohorts**  **(n = 256)** | **Public cohort**  **(*n* = 15)** |
| --- | --- | --- | --- | --- |
| **Manufacturer,**  ***n* (%)** | GE | 398 (74%) | 45 (17%) | — |
|  | Siemens | 112 (21%) | 121 (47%) | — |
|  | Philips | 27 (5%) | 90 (35%) | 15 (100%) |
|  | Others | — | 2 (<2%) | — |
| **Magnetic field strength, *n* (%)** | 1.5 T | 12 (2%) | 52 (20%) | 15 (100%) |
|  | 3 T | 525 (98%) | 204 (79%) | — |
| **Pixel spacing (mm)** | Mean ± SD | 0.36 ± 0.09 | 0.40 ± 0.12 | 0.40 ± 0.03 |
| **Slice thickness (mm), *n* (%)** | 3.0 | 516 (96%) | 217 (85%) | 14 (92%) |
|  | 3.5–4.0 | 21 (4%) | 39 (15%) | 1 (8%) |
| **Image size (px)** | Mode (range) | 512×512 (mode); 384×384 –320×320 | 512×512 (mode); 240×240 –1024×1024 | 400×400 |
| **Number of slices per study** | Mean ± SD | 27 ± 3 | 29 ± 4 | 32 ± 6 |
| **With gland segmentation** | Mean ± SD | 18 ± 3 | 13 ± 3 | 13 ± 5 |
| **With NVB segmentation** | Mean ± SD | 10 ± 4 | 12 ± 3 | 3 ± 2 |

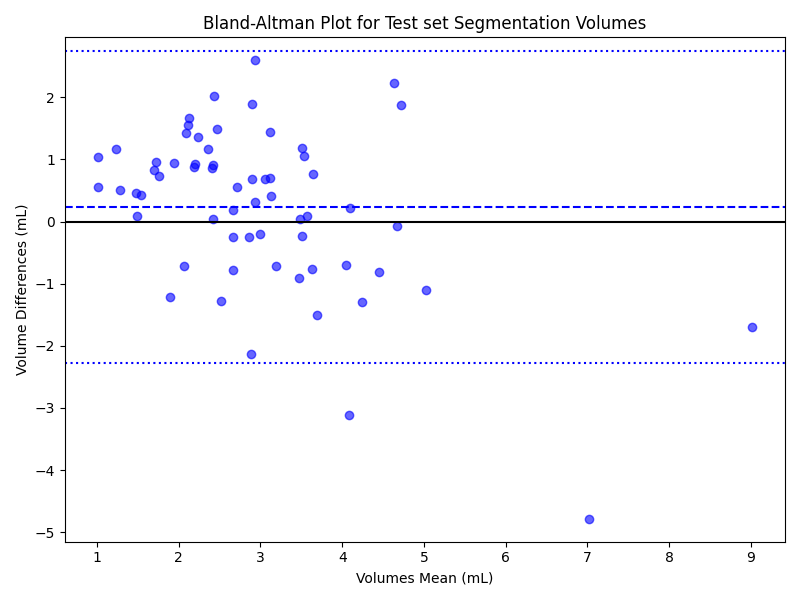

**Figure S2**. Bland-Altman plot for NVB segmentation volumes in the test set. The solid line indicates the mean difference (bias) and the dotted lines denote the 95% limits of agreement. NVB: neurovascular bundle

**Table S3**. Model performance (DSC, ASD) across clinical subgroups: prostatic hyperplasia, high-risk NVB invasion, and peripheral zone lesions. ASD, average surface distance; DSC, Dice similarity coefficient; NVB: neurovascular bundles.

| **Metric / Statistical test** | **Hyperplasia** | **High risk of NVB invasion** | **Peripheral zone lesion** |
| --- | --- | --- | --- |
| **DSC** | | | |
| Normality | *p =* 0.3482 / 0.0994 → *normal* | *p* = 0.3752 / 0.4058 → *normal* | *p* = 0.0406 / 0.2509 → *non-normal / normal* |
| Levene’s test | *p* = 0.0153 → *unequal variances* | *p* = 0.2872 → *equal variances* | *p* = 0.5652 → *equal variances* |
| Selected test | t-test (unequal variances) | t-test | Mann–Whitney U |
| Test result | t = 1.5853, *p* = 0.1183 → *no significant* | t = -1.8063, *p* = 0.0859 → *no significant* | U = 453.0, *p* = 0.8755 → *no significant* |
| Effect size | *d* = 0.37 (*moderate*) | *d* = -0.92 (*large*) | *r* = 0.02 (*small*) |
| **ASD (mm)** | | | |
| Normality (Group 1 / 2) | *p* = 0.0002 / 0.0000 → *non-normal* | *p* = 0.0564 / 0.0306 → *approx. normal / non-normal* | *p* = 0.0000 / 0.0000 → *non-normal* |
| Levene’s test | *p* = 0.1696 → *equal variances* | *p* = 0.2703 → *equal variances* | *p* = 0.2113 → *equal variances* |
| Selected test | Mann–Whitney U | Mann–Whitney U | Mann–Whitney U |
| Test result | U = 446.0, p = 0.9355 → *no significant* | U = 60.0, p = 0.1829 → *no significant* | U = 401.0, p = 0.5457 → *no significant* |
| Effect size | *r* = 0.01 (*small*) | *r* = 0.29 (*moderate*) | *r* = 0.08 (*small*) |

**Table S4**. Comparative metrics of automatic NVB segmentation: Teunissen et. al vs. this study. Values are median (IQR). Teunissen et. al values correspond to the main study (15 patients) and report inter-rater manual segmentation metrics; This study reports the independent test set (*N* = 94) comparing the automatic model with manual reference contours. NVB, neurovascular bundles.

| **Metric** | Teunissen et. al**—Left NVB** | Teunissen et. al **—Right NVB** | **This study— Left NVB** | **This study—Right NVB** |
| --- | --- | --- | --- | --- |
| **Total volume (cc)** | 6.18 (5.22–8.31) | 7.19 (5.83–9.08) | 1.50 (1.14–1.96) | 1.37 (0.98–1.75) |
| **Superior–inferior extent (mm)** | 46.00 (43.00–51.00) | 48.00 (43.00–51.00) | 14.78 (11.91–16.17) | 14.63 (12.03–16.27) |
| **Dice similarity coefficient** | 0.60 (0.54–0.68) | 0.61 (0.53–0.69) | 0.59 (0.49–0.67) | 0.61 (0.51–0.68) |
| **Mean surface distance (mm)** | 1.96 (1.59–2.31) | 1.86 (1.53–2.52) | 1.09 (0.60–2.26) | 0.81 (0.46–1.62) |
| **Hausdorff distance (mm)** | 12.13 (9.36–15.10) | 11.78 (9.52–14.22) | 13.81 (9.90–18.95) | 13.47 (9.49–17.91) |
